# Design, rationale, and baseline characteristics of the SONIC-HF multicenter registry

**DOI:** 10.1101/2024.10.16.24315221

**Authors:** Nobuyuki Kagiyama, Kentaro Kamiya, Misako Toki, Hiroshi Saito, Kentaro Iwata, Yuya Matsue, Kenji Yoshioka, Kazuya Saito, Azusa Murata, Akihiro Hayashida, Junya Ako, Takeshi Kitai, Emi Maekawa

## Abstract

**Background:** Skeletal muscle mass and function are crucial for assessing physical frailty, sarcopenia, and cachexia, which significantly impact the prognosis of geriatric patients with heart failure (HF). Ultrasound-based assessment of skeletal muscles offers a non-invasive, real-time alternative to traditional methods. The *compariSON of various methods In evaluatIon of sarCopenia in patients with Heart Failure* study (SONIC-HF) aimed to evaluate the feasibility and prognostic impact of ultrasound-based muscle assessment in geriatric patients with HF.

**Methods:** This multicenter, prospective cohort study enrolled HF patients aged ≥65 years who could ambulate independently at discharge. Certified observers assessed muscle thickness (biceps, quadriceps, rectus femoris, and diaphragm) using ultrasound at rest and during contraction. The primary endpoint was all-cause mortality. Secondary endpoints included HF hospitalization, unplanned hospital visits, and cardiovascular and non-cardiovascular mortality.

**Results:** Of the 692 enrolled patients (median age 81 (IQR 74–86) years, 57.6% female, left ventricular ejection fraction 45% (32%–60%)), ultrasound-based muscle assessments were completed in 606 patients. Interobserver reliability was excellent (intraclass correlation coefficient 0.84–0.99). Median muscle thicknesses at rest and during contraction were: diaphragm 1.9 (1.6–2.3) mm and 2.9 (2.3–3.8) mm; biceps 19.6 (15.9–23.1) mm and 25.3 (21.3–29.5) mm; quadriceps 19.0 (15.0–23.5) mm and 24.8 (19.9–29.5) mm; rectus femoris 9.7 (7.1–12.3) mm and 12.1 (9.6–15.0) mm. The median follow-up time was 733.5 (438–882) days.

**Conclusions:** The SONIC-HF registry will provide valuable insights into the feasibility and prognostic implications of ultrasound-based muscle assessment in geriatric patients with HF.

## Introduction

Heart failure (HF) is a prevalent and severe condition affecting approximately 26 million people worldwide ^1^. Due to the global aging trend, the number of HF patients continues to rise. Notably, the population aged 65 and over is growing faster than other age groups. By 2050, one in six people globally will be over age 65 (16%), a rise from one in 11 in 2019 (9%) ^2^.

Muscle weakness is a recognized problem in both the general population and HF patients. HF itself predisposes patients to muscle weakness due to factors like inflammation, inactivity, oxidative stress, malnutrition, and muscle cell death ^3^. In our previous research called FRAGILE-HF (prevalence and prognostic value of physical and social frailty in geriatric patients hospitalized for heart failure), we showed that physical frailty ^4, 5^, sarcopenia ^6^, and cachexia ^2, 7^ were highly prevalent among geriatric HF patients and were significantly associated with increased adverse events. The assessments of muscle mass and function in FRAGILE-HF relied on tools like questionnaires, body and limb measurements, handgrip strength tests, and bioelectrical impedance analysis. However, these traditional methods have limitations, particularly in HF patients with edema or other conditions that can obscure accurate assessments of muscle mass.

Ultrasound has emerged as a valuable tool for assessing skeletal muscle mass and function due to its non-invasive nature, real-time imaging capability, and accessibility. Compared with X-rays or magnetic resonance imaging (MRI), ultrasound is more practical for repeated assessments, particularly at the bedside. Additionally, ultrasound enables measurements of muscle thickness both at rest and during contraction, allowing for a dynamic evaluation of muscle function. Despite these advantages, its feasibility and prognostic value have not been extensively studied for patients with HF in multicenter settings.

To address this gap, the SONIC-HF study (*compariSON of various methods In evaluatIon of sarCopenia in patients with Heart Failure*) was initiated to evaluate the feasibility and prognostic impact of ultrasound-based skeletal muscle assessments in this high-risk population. In this paper, we summarize the design and rationale of the study and demonstrate the baseline characteristics of enrolled patients.

## Methods

### Study design

The SONIC-HF study was a multicenter, prospective cohort study conducted across four hospitals in Japan, aimed at evaluating the prevalence of muscle atrophy and its prognostic implications in geriatric HF patients. The study primarily assessed muscle mass and function using ultrasound, supplemented by anthropometric measurements. The protocol followed the Declaration of Helsinki and received approval from the institutional review boards of all participating centers. All participants were fully informed about the study’s objectives and procedures through an opt-out method and were assured of their right to withdraw at any time. As this was an observational study that did not entail invasive procedures or interventions, written informed consent was not required under the Ethical Guidelines for Medical and Health Research Involving Human Subjects, as per the Japanese Ministry of Health, Labour and Welfare.

Comprehensive details of the study, including its objectives, eligibility criteria, primary outcomes, and participating hospitals, were registered and made publicly accessible on the University Hospital Information Network (UMIN-CTR, unique identifier: UMIN000031635) before the enrollment of the first participant. Although the study originally planned to involve six hospitals, logistical challenges prevented two centers from enrolling patients.

Eligible patients were those aged 65 years or older, admitted for decompensated HF, and able to ambulate independently at discharge. The study included only the first hospitalization for each patient during the study period, with HF decompensation diagnosed according to the Framingham criteria ^8^. Patients were excluded if they had undergone heart transplantation, were using a left ventricular assist device, were receiving chronic peritoneal dialysis or hemodialysis, or were diagnosed with acute myocarditis. Additionally, patients were excluded if they had low B-type natriuretic peptide (BNP) levels, lacked BNP or N-terminal-proBNP (NT-proBNP) data, or had BNP levels below 100 pg/mL or NT-proBNP levels below 300 pg/mL at admission. The study cohort included patients with both HF with reduced and preserved ejection fraction.

### Data acquisition

Patient characteristics, including demographic and laboratory data, physical function, and ultrasound-based muscle measurements, were collected prospectively once the patient had stabilized following the completion of intravenous drug administration. Echocardiography was also performed before discharge, in accordance with published guidelines ^9^. Significant valvular heart diseases were defined as moderate or severe valvular heart disease ^10^.

To assess muscle thickness, specific anatomical sites were evaluated. The midpoint between the acromion and olecranon was used to measure the thickness of the biceps brachii and brachialis muscles, with the patient either seated in a chair or lying on a bed (Figure 1, Panel A). The thickness of the quadriceps and rectus femoris muscles was measured at the midpoint between the lateral condyle and the greater trochanter, with the patient in a supine position (Figure 1, Panel B). A linear probe with a frequency range of 7.5 – 13.0 MHz was placed perpendicular to the tissue interface, and the distance from the adipose tissue-muscle interface to the muscle-bone interface was recorded as muscle thickness ^11, 12^. Muscle contraction was evaluated using isotonic and isometric exercises, with the upper limbs assessed during arm bending and the lower limbs during leg extension without bending ^11, 12^. Diaphragm thickness was measured between the right anterior and mid-axillary lines at the lowest intercostal space using the same probe as for the upper and lower limb measurements (Figure 1, Panel C) ^13^. These measurements were performed twice during both natural expiration (at rest) and full inspiration, with the mean value of the two measurements used for analysis.

**Figure 1.**
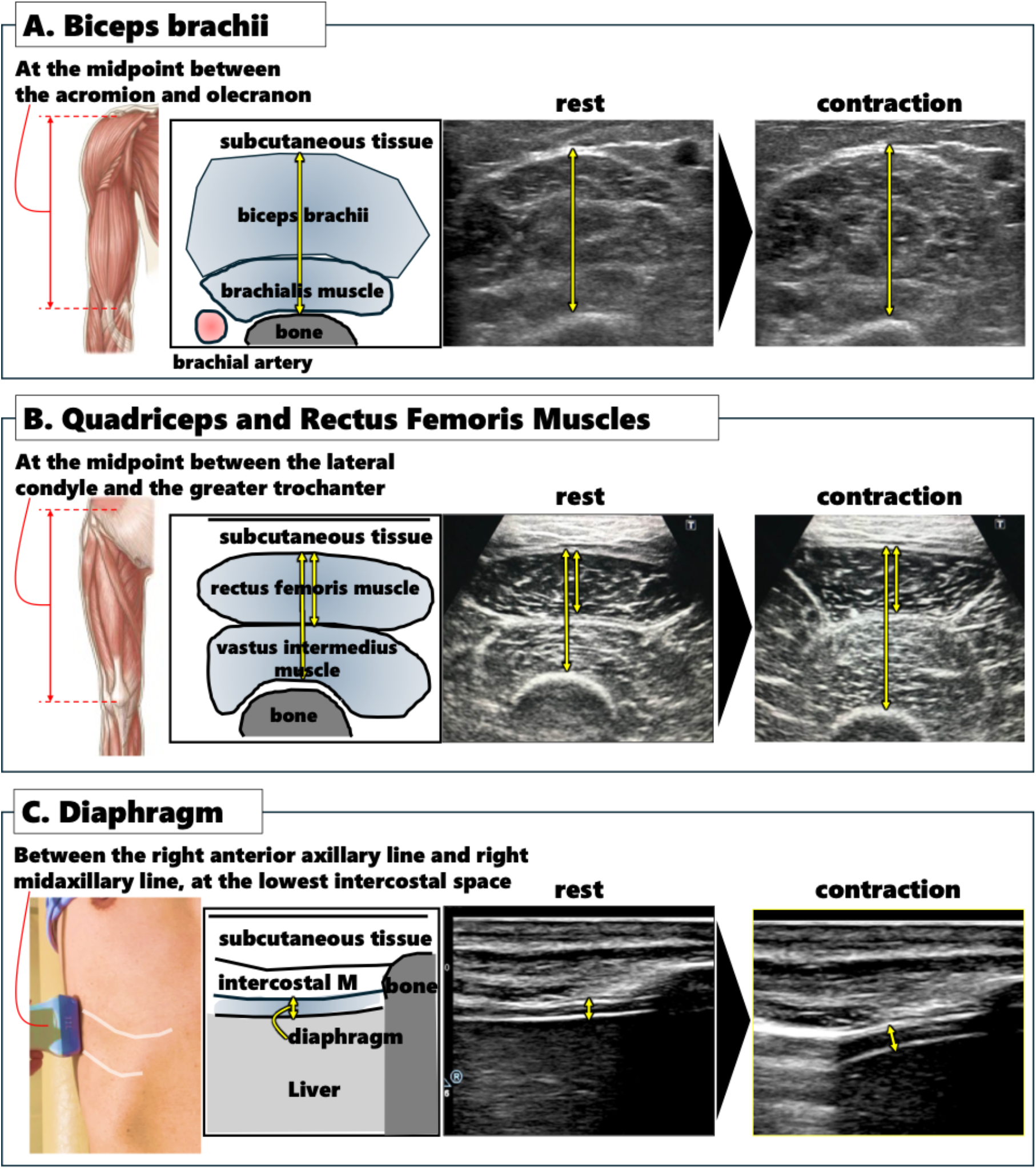
Ultrasound assessment of skeletal muscles. Panel A: Measurement of the biceps brachii and brachialis thickness at the midpoint between the acromion and olecranon, with the patient seated or lying down. Panel B: Quadriceps and rectus femoris muscle thickness measured at the midpoint between the lateral condyle and the greater trochanter, with the patient in a supine position. Panel C: Diaphragm thickness measured between the right anterior and mid-axillary lines at the lowest intercostal space, during both natural expiration and full inspiration.

Since many observers were initially unfamiliar with the measurement procedure, a certification program for ultrasound scanning was implemented. Observers practiced scanning on volunteers and submitted images to the core lab for feedback. Only after the core lab certified that the observer could reliably measure muscles without significant errors, they were permitted to scan actual patients. All measurements were conducted twice, both at rest and during contraction, with the mean value of the two measurements used for analysis.

Interobserver variability was assessed by two blinded observers who independently measured the same randomly selected images twice. The number of images used for this analysis ranged from 40 to 46 for each muscle and phase. The average values from each observer were compared, and the level of agreement between them was determined to evaluate interobserver reliability.

### End points

The primary outcome predefined in this study was all-cause mortality. After discharge, most patients had follow-up appointments at outpatient clinics at least every three months, with additional visits as required by their individual medical needs. For patients who did not attend these clinics, outcome data were obtained via telephone interviews, either by reviewing medical records from other healthcare facilities or by speaking with the patient’s family. Causes of death were categorized as either cardiovascular or non-cardiovascular, following established definitions. Cardiovascular deaths included those resulting from HF, acute coronary syndrome, stroke, ventricular arrhythmias, and other cardiovascular diseases. Sudden death from an unknown cause was also classified as cardiovascular death ^14^.

### Statistical analysis

Continuous variables are presented as mean ± standard deviation or as medians with interquartile ranges (IQR) (1st and 3rd quartiles), depending on their distribution. Categorical variables are expressed as frequencies and percentages. To adjust for the association between the variable of interest and outcomes, including all-cause mortality and the composite of HF hospitalization and all-cause mortality, we employed the Meta-analysis Global Group in Chronic Heart Failure (MAGGIC) risk score ^15^ and BNP levels as covariates. The MAGGIC risk score has been well-validated for predicting outcomes in Japanese patients with HF, ^16, 17^, and the inclusion of BNP levels at discharge has been shown to improve the score’s discrimination while maintaining adequate calibration ^17^.

## Results

### Population

A total of 692 patients were enrolled in the SONIC-HF study across four participating hospitals in Japan. Table 1 summarizes the patient characteristics. The median age of the cohort was 81 years (74 – 86 years), with 42.4% male and 57.6% female. The median body mass index was 21.1 kg/m^2^ (19.0 – 23.7 kg/m^2^), and the median body surface area was 1.51 m^2^ (1.37 – 1.64 m^2^). The median systolic blood pressure at admission was 112.0 mmHg (102.0 – 124.8 mmHg), and the median diastolic blood pressure was 61.0 mmHg (56.0 – 68.0 mmHg). The median heart rate was 70 bpm (61 – 80 bpm). Regarding the New York Heart Association (NYHA) classification, 19.9% of the patients were classified as NYHA class III-IV, indicating a significant proportion of patients with advanced symptoms even after completion of intensive HF management.

**Table 1.**
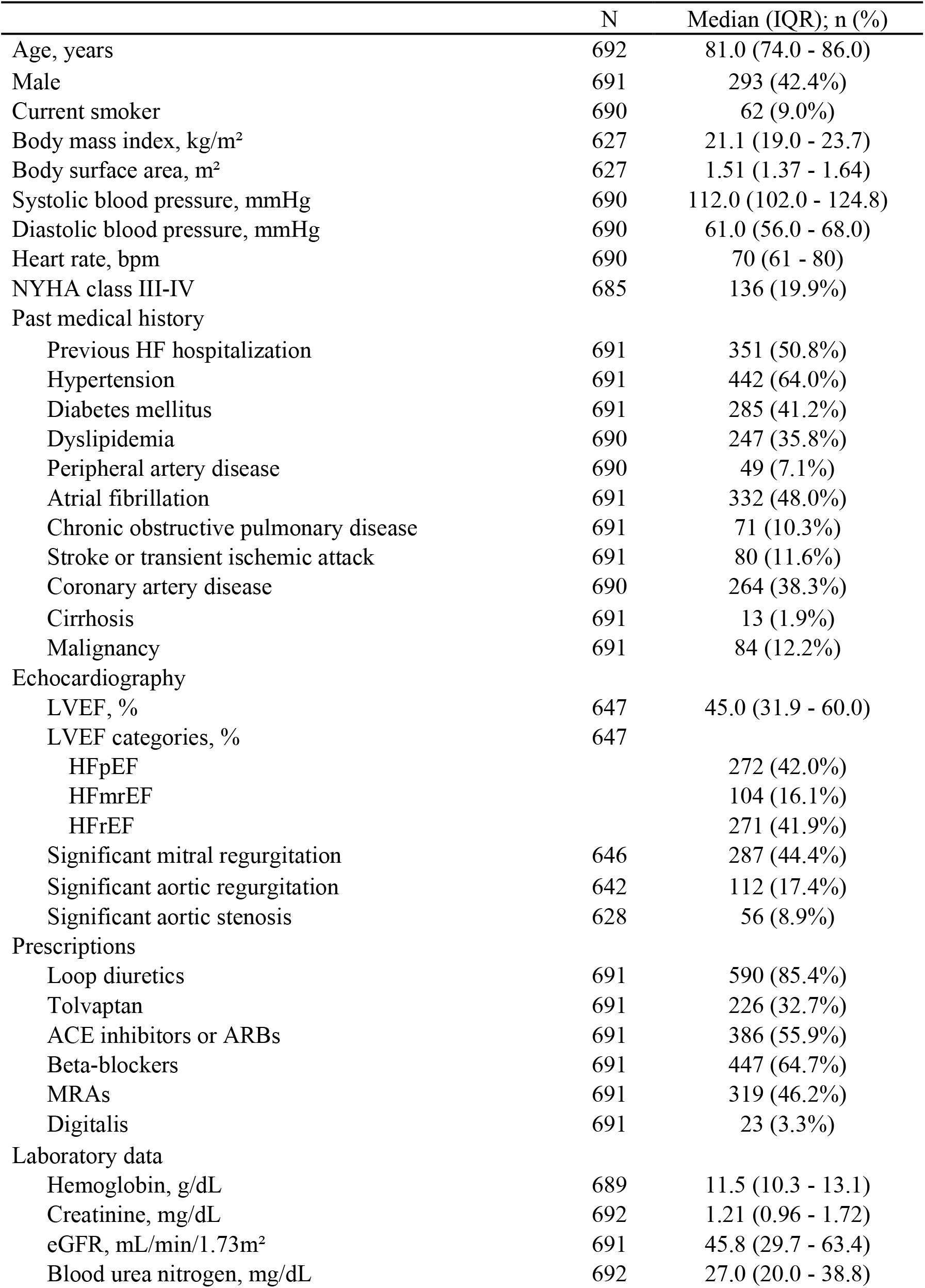
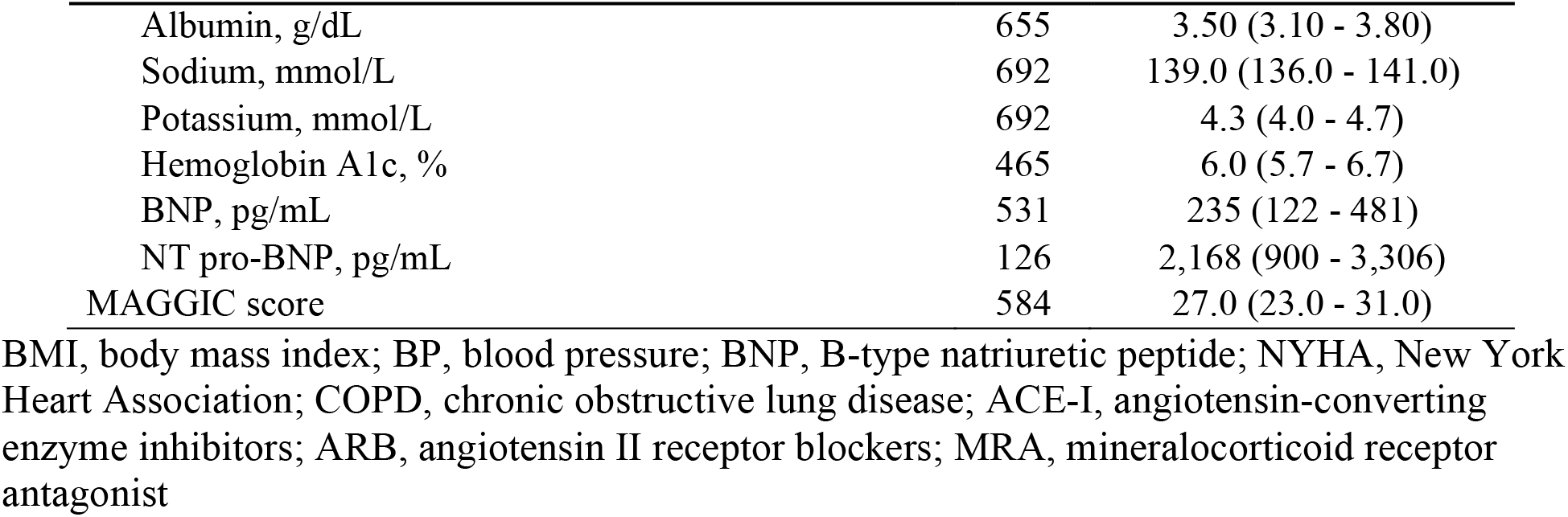
Patient characteristics.

The cohort exhibited a high prevalence of comorbid conditions, which is typical for this demographic. A previous history of HF hospitalization was present in 50.8% of patients, and hypertension was the most common comorbidity, affecting 64.0% of the cohort, followed by atrial fibrillation (48.0%), diabetes mellitus (41.2%), and dyslipidemia (35.8%). Other notable comorbidities included coronary artery disease (38.3%), stroke or transient ischemic attack (11.6%), and chronic obstructive pulmonary disease (10.3%).

Echocardiographic assessments revealed a median left ventricular ejection fraction (LVEF) of 45.0% (31.9% – 60.0%). The distribution of LVEF categories was relatively balanced: 42.0% of patients had HF with preserved ejection fraction (HFpEF), 41.9% had HF with reduced ejection fraction (HFrEF), and 16.1% had HF with mildly reduced ejection fraction (HFmrEF). Additionally, significant mitral regurgitation was present in 44.4% of patients, while significant aortic regurgitation and aortic stenosis were observed in 17.4% and 8.9% of patients, respectively.

In terms of pharmacotherapy and laboratory data, loop diuretics were prescribed to 85.4% of the cohort. Other frequently prescribed medications included beta-blockers (64.7%), angiotensin-converting enzyme inhibitors or angiotensin II receptor blockers (55.9%), and mineralocorticoid receptor antagonists (46.2%). The median hemoglobin level was 11.5 g/dL (10.3 – 13.1 g/dL), and the median creatinine level was 1.21 mg/dL (0.96 – 1.72 mg/dL), resulting in an estimated glomerular filtration rate of 45.8 mL/min/1.73 m^2^ (29.7 – 63.4 mL/min/1.73 m^2^). The median BNP level was 235 pg/mL (122 – 481 pg/mL), and the median NT-proBNP level was 2,168 pg/mL (900 – 3,306 pg/mL). The MAGGIC risk score had a median value of 27.0 (23.0 – 31.0).

### Ultrasound-based muscle assessment

Ultrasound-based skeletal muscle assessments were successfully conducted in 606 of the 692 patients initially enrolled in the SONIC-HF study. The difference in sample size was primarily due to patients being discharged early or unexpectedly, before completing the ultrasound examinations.

Interobserver variability was rigorously assessed to ensure the reliability of the ultrasound measurements, with intraclass correlation coefficients (ICCs) calculated for each muscle group. The biceps brachii showed an ICC of 0.989 (0.978 – 0.995) at rest, indicating high consistency, and 0.973 (0.948 – 0.986) during contraction. Similarly, the quadriceps demonstrated an ICC of 0.979 (0.963 – 0.988) at rest and 0.997 (0.994 – 0.998) during contraction, suggesting excellent reproducibility. The rectus femoris muscle had slightly lower but still robust ICC values, with 0.883 (0.798 – 0.934) at rest and 0.949 (0.909 – 0.971) during contraction. Diaphragm measurements, while showing lower ICCs, were still within acceptable ranges, with an ICC of 0.805 (0.669 – 0.889) at rest and 0.799 (0.653 – 0.888) during inspiration.

The median muscle thicknesses observed across the cohort are summarized in Table 2. Specifically, the biceps brachii had a thickness of 19.6 mm (15.9 – 23.1 mm) at rest and 25.3 mm (21.3 – 29.5 mm) during contraction. For the quadriceps, the thickness was 9.7 mm (7.1 – 12.3 mm) at rest and increased to 12.1 mm (9.6 – 15.0 mm) during contraction. The rectus femoris had a median thickness of 19.0 mm (15.0 – 23.5 mm) at rest, which increased to 24.8 mm (19.9 – 29.5 mm) during contraction. Finally, the diaphragm, a key respiratory muscle, had a median thickness of 1.88 mm (1.55 – 2.30 mm) at rest and 2.90 mm (2.25 – 3.75 mm) during inspiration.

**Table 2.** Ultrasound-based skeletal muscle thickness.

|  | N | Median (IQR); n (%) |
| --- | --- | --- |
| Bicep thickness at rest, mm | 606 | 19.6 (15.9 – 23.1) |
| Bicep thickness during contraction, mm | 604 | 25.3 (21.3 – 29.5) |
| Rectus femoris thickness at rest, mm | 604 | 9.7 (7.1 – 12.3) |
| Rectus femoris thickness during contraction, mm | 603 | 12.1 (9.6 – 15.0) |
| Quadriceps thickness at rest, mm | 606 | 19.0 (15.0 – 23.5) |
| Quadriceps thickness during contraction, mm | 605 | 24.8 (19.9 – 29.5) |
| Diaphragm thickness at rest, mm | 605 | 1.88 (1.55 – 2.30) |
| Diaphragm thickness during inspiration, mm | 606 | 2.90 (2.25 – 3.75) |

The study has completed its follow-up period, with a median follow-up duration of 733.5 (438.0–882.0) days, and survival analyses are set to be performed.

## Discussion

As the average age of HF patients increases worldwide, comorbidities associated with HF are becoming more prevalent. Physical frailty, sarcopenia, and cachexia are particularly common comorbidities, especially in geriatric HF patients. The SONIC-HF registry focuses on assessing muscle mass, a cornerstone in evaluating physical frailty, sarcopenia, and cachexia in HF patients aged 65 years and older, with the aim of validating simple and widely applicable ultrasound-based techniques.

Mass and function are two key aspects of skeletal muscles and form the core of the pathophysiology of physical frailty and sarcopenia. Physical frailty is most often defined by five components: weight loss, exhaustion, low physical activity, slowness, and weakness ^18^, while sarcopenia is characterized by muscle strength, muscle mass, and physical performance ^19, 20^. Cachexia is defined by weight loss, decreased muscle strength, fatigue, anorexia, low fat-free mass, and abnormal biochemistry ^2, 21^. These comorbidities are common and significantly impact mortality and quality of life in patients with HF, and accordingly, it is highly recommended to assess these conditions in geriatric HF patients (REF). However, in clinical practice, the gold-standard methods for assessing muscle mass and function—dual-energy X-ray absorptiometry, MRI, or computed tomography—are rarely performed. Bioelectrical impedance analysis is easier but lacks sufficient accuracy, particularly in HF patients who tend to have edema and increased extracellular water ^22^. Therefore, there is a strong need for simpler and more widely applicable methods to accurately assess muscle mass and function.

Ultrasound is widely employed in the medical assessment of various organs. It operates by emitting high-frequency sound waves, which are reflected off tissues and processed to generate detailed images. Skeletal muscles are well-suited for ultrasound assessment because they are located near the body’s surface, allowing for clear visualization with minimal interference from air or bone. Additionally, tissues close to the probe are imaged with greater clarity than deeper structures due to reduced signal dissipation. Furthermore, advances in ultrasound technology have led to the development of portable, handheld devices, which are now commonly used in clinical practice and are increasingly regarded as an extension of the physical examination.

Owing to these advantages, ultrasound has been increasingly used in the assessment of skeletal muscles. Sanada et al. validated the accuracy of ultrasound-based muscle mass assessment in the intensive care setting, comparing it with MRI in 72 participants, and found strong correlations between muscle mass measured by MRI and muscle thickness measured by ultrasound 23. Ogawa et al. investigated the reproducibility of ultrasound assessment of quadriceps thickness, reporting similar ICCs of 0.96 – 0.99 ^24^. Watanabe et al. demonstrated that quadriceps muscle thickness decreases with age in healthy Japanese individuals, with mean values of 25.9 ± 5.7 mm for men and 20.7 ± 4.8 mm for women aged 65 years or older ^25^. The median quadriceps thickness in our cohort was 20.1 mm (IQR 19.2 – 20.9 mm) at rest, which was thinner than in the above studies but aligned with previous findings that reported a median value of 20.1 mm (IQR 19.2 – 20.9 mm) in patients with HFpEF ^26^. Other studies have also demonstrated the association of skeletal muscle thickness and exercise capacity in patients with HF ^27-29^. Although these studies establish reliability and associations with muscle mass as measured by gold-standard methods, the prognostic value of ultrasound-based muscle thickness in HF patients remains underexplored. Our multicenter registry will contribute significant insights into the literature regarding the prognostic value of these measurements.

Our study has several limitations. First, this registry does not employ core-laboratory analysis of ultrasound images, and the ultrasound equipment used is not standardized. As a result, ultrasound intensity was not measured in this study. The intensity of ultrasound signals from skeletal muscles may provide valuable information beyond muscle thickness ^30^. However, this requires offline analysis, which is not available on the machines used and significantly limits its clinical application. Therefore, we focused on muscle thickness, which is easier to measure at the bedside. Additionally, the study was conducted at only four sites and included only Japanese patients. Since body surface area and body mass index in Japanese patients differ significantly from those in Western populations, caution is needed when generalizing these findings to Western populations.

## Conclusions

The SONIC-HF registry has included 692 patients with geriatric patients with HF in Japan and measured ultrasound-based skeletal muscle thickness for 606 patients. The study tracked patients for two years and has detailed baseline and follow-up information. This will be the first and largest database of ultrasound-based skeletal muscle thickness for patients with HF, and will uncover the clinical usefulness of this easy, reproducible, and widely-applicable method of muscle assessment for patients with HF.

## Data Availability

All data produced in the present study are available upon reasonable request to the authors.

## Abbreviations

BNP: B-type natriuretic peptide
MRI: magnetic resonance imaging
NYHA: New York Heart Association
HF: heart failure
HR: hazard ratio

